# A Systematic Review of Mendelian Randomization Studies on Celiac Disease

**DOI:** 10.1101/2024.07.03.24309885

**Authors:** Mahmud Omar, Reem Agbareia, Salih Nassar, Mohammad Omar, Mohammad E. Naffaa, Adi Lahat, Kassem Sharif

## Abstract

**Background and Objective:** Mendelian randomization (MR) has become an important tool in epidemiology, used to infer causal relationships diseases. This review aims to consolidate existing MR evidence concerning celiac disease (CeD).

**Methods:** We systematically searched major databases up to May 2024, adhering to PRISMA guidelines. Only MR studies explicitly investigating CeD were included. We assessed the quality of each study based on the strength, independence, and exclusivity of the instrumental variables used.

**Results:** From an initial pool of 207 articles, 35 met our inclusion criteria. These studies frequently addressed the relationship between CeD and autoimmune diseases like inflammatory bowel disease (IBD) and explored connections with gut microbiota, various cancers, and metabolic disorders. Significant findings highlight a robust bi-directional association between IBD and CeD and complex interactions with gut microbiota. Notably, many associations reported were near the threshold of clinical significance.

**Conclusion:** This systematic review highlights the dual nature of current MR evidence on CeD. On one hand, we observe consistent associations between CeD, IBDs, and gut microbiota. On the other, there is a plethora of weaker associations that raise critical questions about their clinical and research significance. This work lays a solid foundation for deeper investigations into these weaker links, particularly in relation to lymphomas and psychiatric conditions. It calls for an expanded use of MR and other methodologies to explore under-researched areas.

## Introduction

Mendelian randomization (MR) has emerged as a powerful epidemiological tool, leveraging genetic variants as instrumental variables to infer causal relationships between modifiable exposures and health outcomes (1,2).

MR is an analytical approach that uses genetic variants as instruments to estimate the causal effect of an exposure on an outcome (2). This method has the power to mitigate confounding and reverse causation, two common issues in observational studies, by leveraging the random allocation of alleles at conception (3). Thus, MR can provide more reliable evidence of a causal association than traditional epidemiological studies (1,2).

The growing accessibility to large-scale genetic data has catalyzed a surge in MR studies, expanding our understanding of the genetic underpinnings of various diseases (4). Among these, celiac disease (CeD) has been a notable focus of investigation (5). Recent studies have traversed a spectrum of findings. Some identified clear non-associations that suggest avenues requiring no further inquiry (6). Other studies established associations that enhance our current understanding, and tentative links that call for deeper exploration (7–9). Yet, some areas remain under-researched, presenting fresh opportunities for novel insights.

Given the robust nature of evidence that MR provides, this review aims to synthesize and organize the existing MR literature on celiac disease. By doing so, we intend not only to consolidate current knowledge but also to guide future research directions in this evolving field. This systematic approach seeks to clarify the genetic contributions to celiac disease, delineating well-supported findings from those warranting further investigation.

## Materials and Methods

### Registration and Protocol

This systematic review was registered with the International Prospective Register of Systematic Reviews (PROSPERO) under the registration code CRD42024545782 (10). Our approach adheres to the Preferred Reporting Items for Systematic Reviews and Meta-Analyses (PRISMA) guidelines (11).

### Search Strategy

We executed a search across five major databases: PubMed, Embase, Web of Science, Scopus, and the Cochrane Library, covering publications up to May 2024. We employed database-specific Boolean search strings, which are detailed in the **Supplementary Materials**. To enhance our search, we also conducted manual reference checks of included studies and targeted searches using Google Scholar.

### Study screening and selection

We included only Mendelian randomization studies. Exclusion criteria were set to omit studies not employing Mendelian randomization analysis, studies not specifically analyzing celiac disease, preprints, review articles, case reports, commentaries, protocols, editorials, and non-English publications. Initial screening was facilitated by the Rayyan web application, with two reviewers (MO and KS) independently conducting the selection based on pre-defined criteria (12). Disagreements were resolved by discussion, and inter-rater reliability was quantified using Fleiss’ kappa (13).

### Data Extraction

Data were systematically extracted by MO and KS using a standardized form to capture key details including author names, publication year, exposure and outcome variables, data sources, number of Single Nucleotide Polymorphisms (SNPs), and any conducted sensitivity analyses. Discrepancies were addressed through discussion, with a third reviewer available for consultation if needed.

### Risk of Bias Assessment

We adopted a risk of bias assessment method from Luo et al. (2022) due to the absence of specific tools for MR studies (14). This method focuses on three crucial instrumental variable (IV) assumptions necessary for valid causal inference:

1. <u>IV1: Relevance</u> - Evaluated the strength of association between genetic variants and the exposure. IV1 was considered high if variants demonstrated genome-wide significant P-values (<5×10^–8) and an F-statistic >10, medium for less stringent P-values, and low if these criteria were unmet.
2. <u>IV2: Independence</u> - Assessed the independence of genetic variants from potential confounders using individual-level data or curated databases. IV2 was rated high for robust evaluations, medium for descriptive assessments, and low if not addressed.
3. <u>IV3: Exclusion-Restriction</u> - Investigated indirect genetic effects through various statistical methods. IV3 was deemed high for thorough evaluations, medium if only described, and low if ignored.

Each study was independently reviewed by two investigators, with any discrepancies resolved through consensus.

## Results

### Search Results and Study Selection

Our search strategy initially identified 207 articles. After eliminating 144 duplicates, 63 articles remained for title and abstract screening. Title and abstract screening excluded 17 articles, leaving 46 for full-text review. Following a comprehensive review, 35 studies met the inclusion criteria (5–9,15–43). The inter-rater reliability, assessed by Fleiss’ kappa, was at 0.779, indicating a substantial agreement between the two reviewers (13).

The selection process is depicted in a PRISMA flowchart included in **Figure 1**.

**Figure 1:**
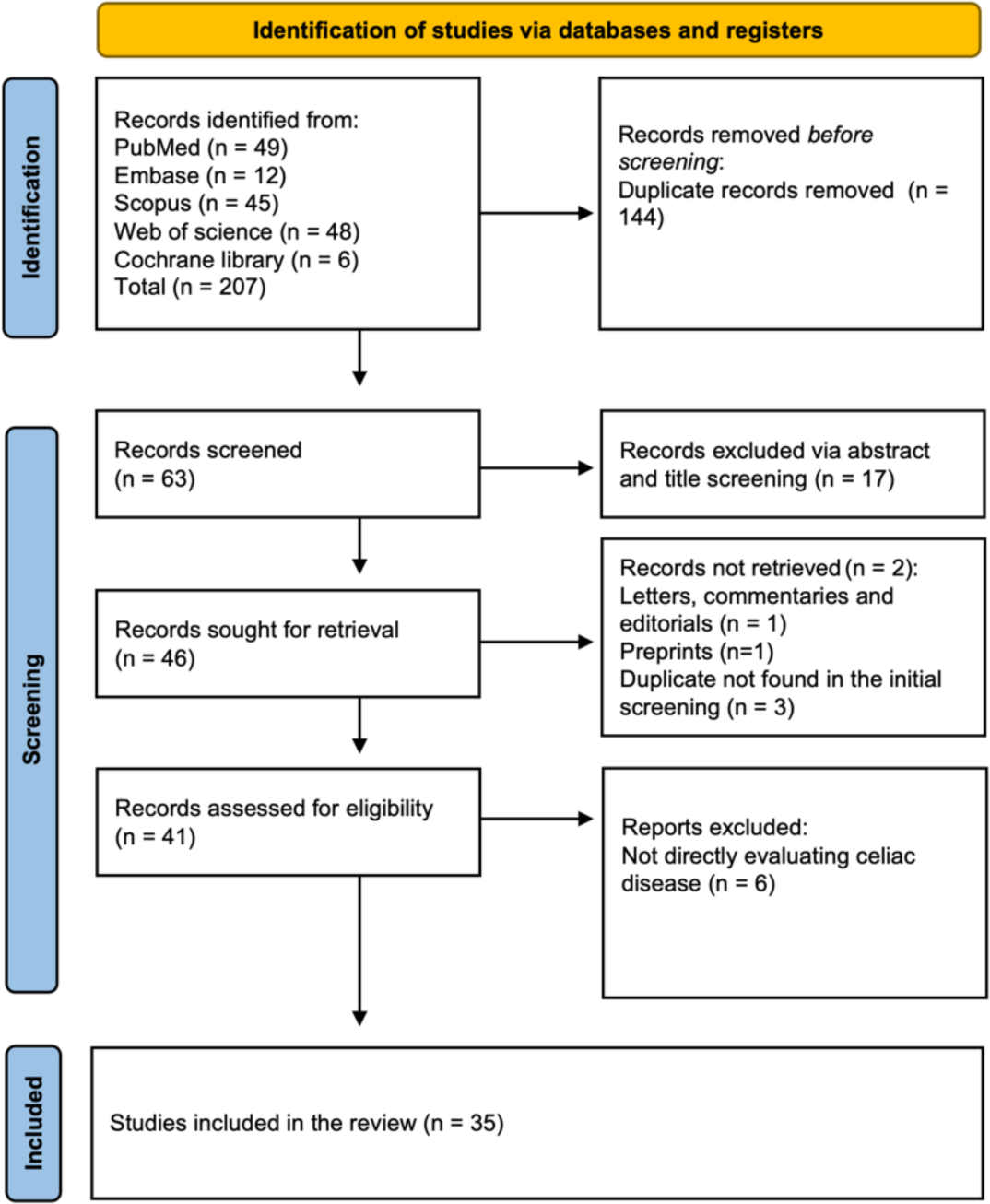
PRISMA flowchart.

### Risk of bias assessment

Most of the included studies were rated as high quality across all three IVs, with 24 studies achieving this standard. Six studies received a moderate rating for one of the IVs but were rated high on the others. The remaining studies had at least one poor rating. IV2 (Independence) had the highest number of non-high ratings, all of which were moderate because these studies only described the assumption without thorough evaluation. The results of the risk of bias assessment are detailed in **Table S1** in the **Supplementary Materials**.

### An overview of the included studies

Our systematic review encompassed 35 studies published between 2022 and 2024, utilizing Mendelian randomization to investigate the genetic associations with CeD. These studies covered a range of health-related categories: 12 addressed autoimmune and inflammatory diseases, four focused on cancer, four examined metabolites and gut microbiota, and 15 explored various other health comorbidities including psychiatric conditions like PTSD and depression, cardiovascular diseases, infections, and more (**Figure 2**). Celiac disease was the primary exposure in 21 studies, while the remaining 14 investigated it as an outcome. The data sources were diverse, predominantly GWAS databases, along with the UK Biobank and FinnGen, and others such as the COVID-19 Host Genetics Initiative and MRC Integrative Epidemiology Unit (MRC-IEU). The studies analyzed a broad range of SNPs, with some examining as few as three to five SNPs and others up to 87 to 97 SNPs. Methodologically, the studies primarily employed the Inverse Variance Weighted (IVW) method, often supplemented with MR-Egger and Weighted Median methods.

**Figure 2:**
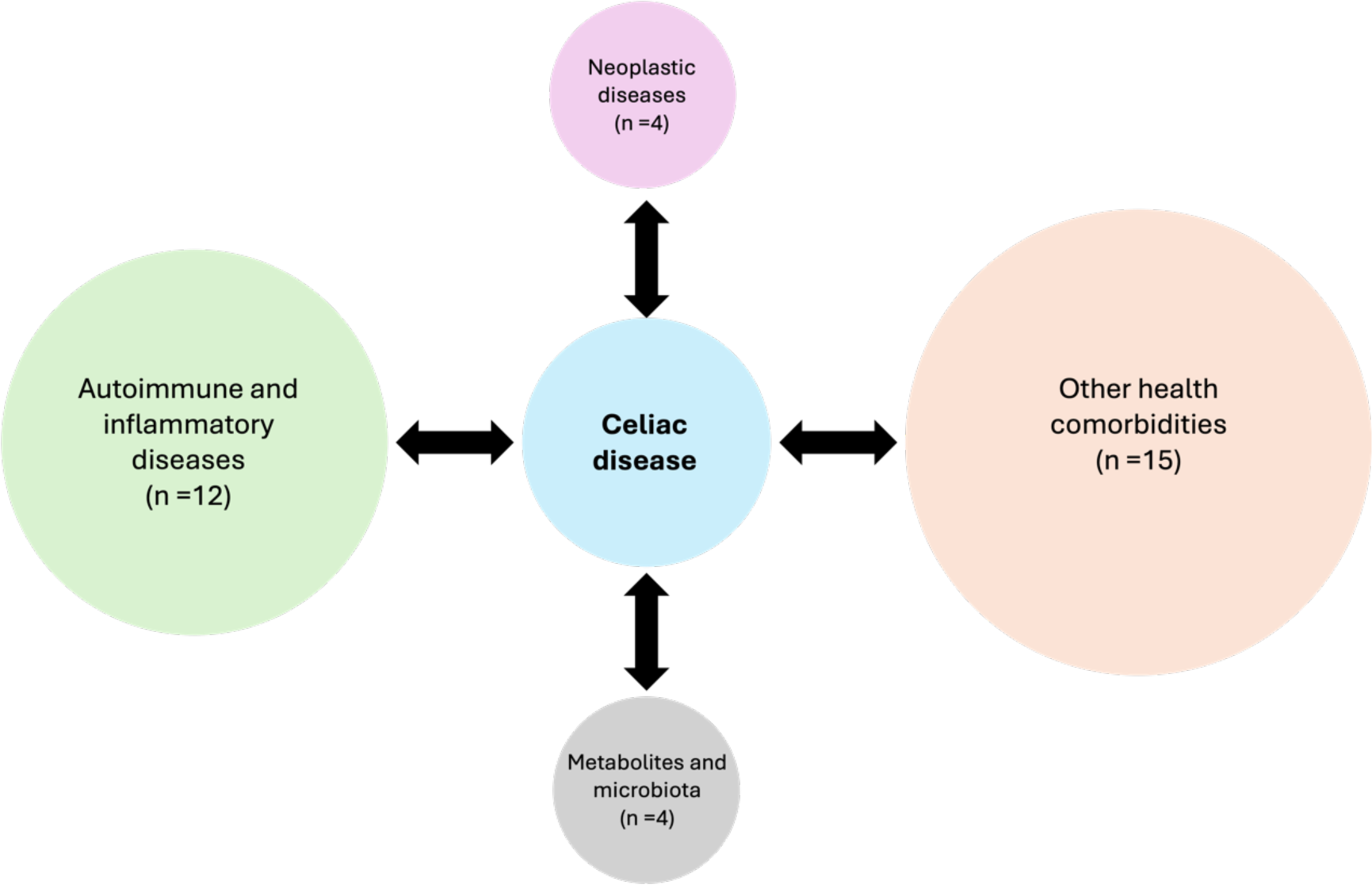
The broad categorization of the studied associations.

Most studies conducted various sensitivity analyses, predominantly MR-PRESSO and leave-one-out tests. Additionally, 19 studies also performed reverse analyses to further validate their findings. A detailed description of the included studies is shown in **Table 1**.

### Autoimmune and Inflammatory Diseases

#### Inflammatory Bowel Disease (IBD)

Four studies have investigated the genetic connections between CeD and IBD, revealing a robust bidirectional relationship. Yuan et al. (2023) noted a significant genetic susceptibility where CeD was linked to an increased risk of IBD, especially Crohn’s disease (OR = 1.1865, P < 0.01), suggesting a shared pathogenetic pathway. Shi et al. (2022) observed a strong bidirectional influence, with a particularly pronounced causal link from Crohn’s disease to CeD (IVW OR = 1.27, 95% CI = 1.19–1.35, P < 0.01). Zhou et al. (2) (2024) and Gu et al. (2022) supported these findings, indicating that genetic factors associated with IBD significantly increase the risk of developing CeD (OR = 1.14, 95% CI = 1.03–1.25, P = 0.01; OR = 1.0828, 95% CI = 1.0258–1.1428, P < 0.01) and vice versa (**Figures 3,4**).

**Figure 3:**
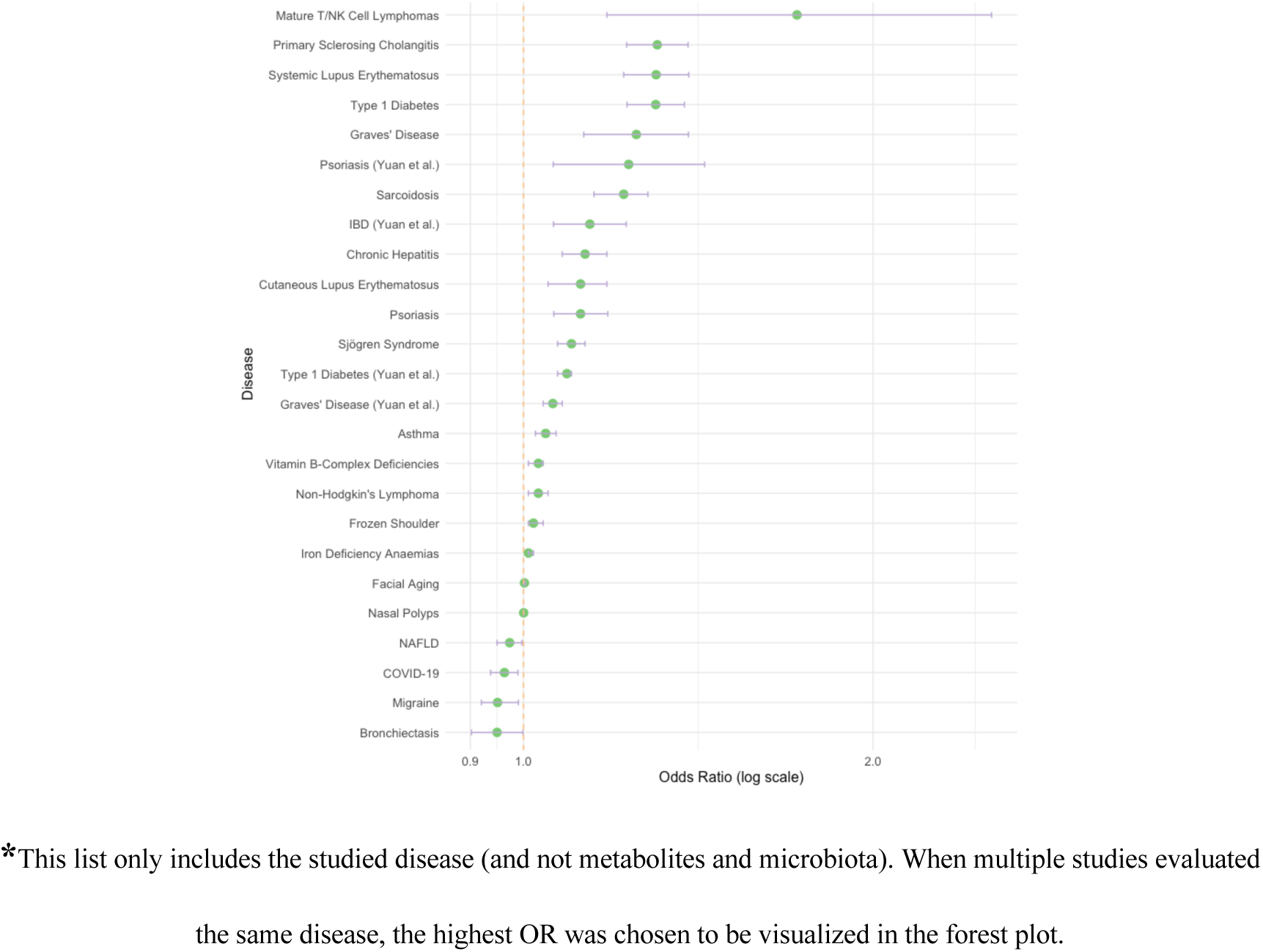
A forest of the associations for CeD as an exposure.

**Figure 4:**
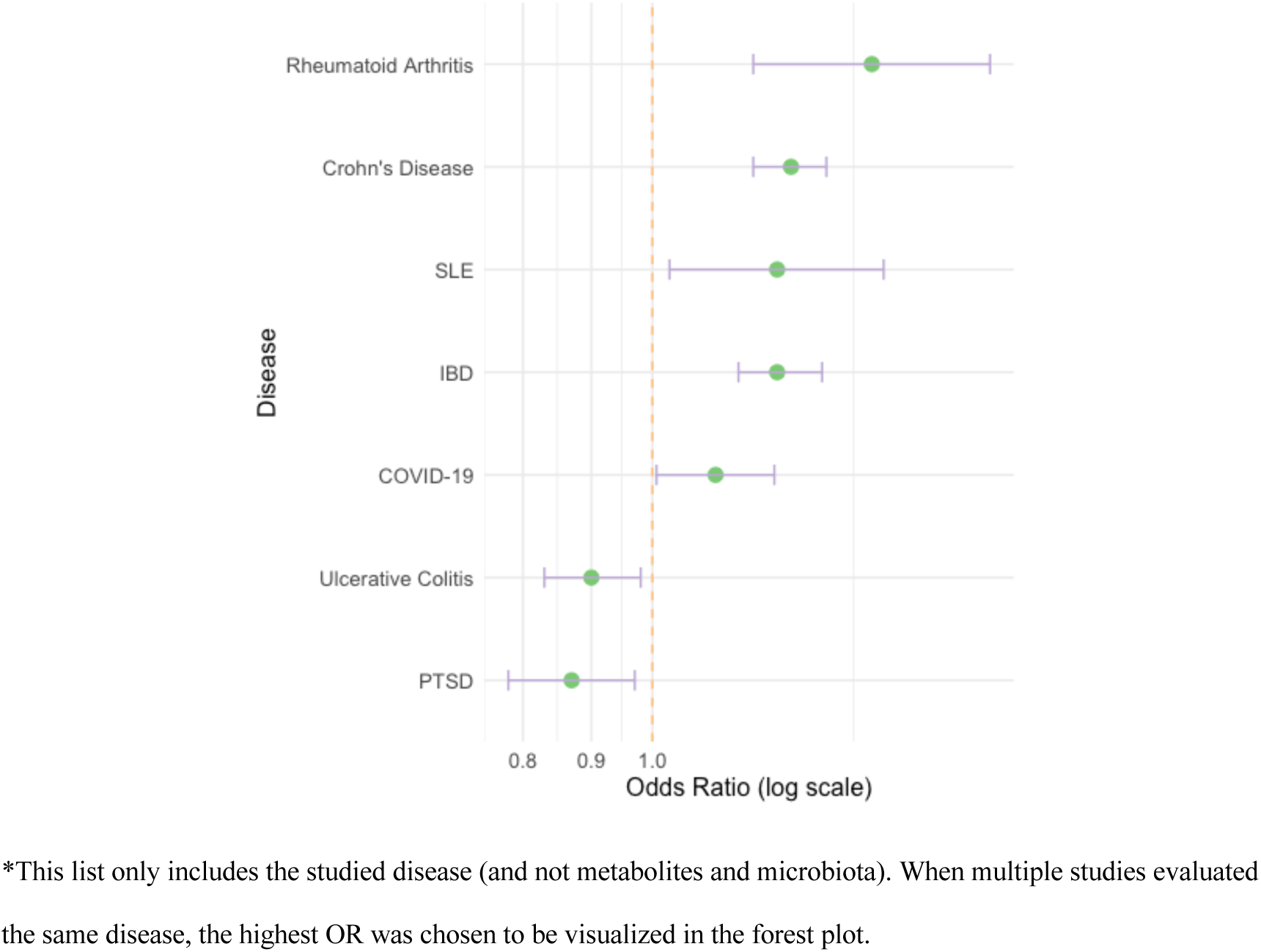
A forest of the associations for CeD as an outcome.

#### Sarcoidosis

Two studies investigated this association. Sun et al. (2024) identified a significant correlation where genetic markers of CeD were associated with an increased risk of developing sarcoidosis (OR = 1.22, 95% CI = 1.15 to 1.28, P < 0.01). Zhou et al. (1) (2024) confirmed this, providing evidence of a causal relationship between CeD and an increased risk of sarcoidosis (IVW OR = 1.13, 95% CI: 1.07–1.20, P < 0.01) (**Figures 3,4**).

#### Other Autoimmune and Inflammatory Diseases

Li et al. (2023) expanded these insights to include a wider range of autoimmune diseases. Their comprehensive analysis showed that CeD increases the risk of diseases such as Crohn’s disease, Graves’ disease, and systemic lupus erythematosus, potentially exacerbating their pathogenesis. Similarly, Wang et al. (2) (2023) investigated the association between SLE and CeD, finding a significant risk increase when SLE was the exposure (OR = 1.70, 95% CI = 1.16–2.50, P < 0.01). Additional studies by Su et al. (2024) and Hua et al. (2022) identified both protective and risk-enhancing genetic influences of CeD. Su et al. observed a protective effect against bronchiectasis (OR = 0.949, P = 0.044), while Hua et al. reported a causal effect of rheumatoid arthritis increasing the risk of CeD (OR = 1.46, P < 0.01). Li et al. (5) found that genetic predispositions to CeD could increase the risk of psoriasis (OR = 1.232 [1.061–1.432], P < 0.01).

Interestingly, Chen et al. discovered a gender-specific association of CeD with nasal polyps in females (OR = 1.000494, 95% CI = 1.000067-1.000922) (**Figures 3,4**). Notably, Li et al. reported non-significant associations for genetic predispositions for CeD with UC, autoimmune hypothyroidism, multiple sclerosis, psoriasis, and ankylosing spondylitis.

### Metabolites and gut microbiota

Four studies evaluated the genetic interplay between CeD and different macrobiotic species and metabolites.

Wang et al. (2024) discovered a significant causal link between elevated serum levels of 1-oleoylglycerophosphoethanolamine and an increased susceptibility to CeD, with an odds ratio (OR) of 11.271 (95% CI: 2.053-61.882, P = 0.005). This points to specific metabolites as potential biomarkers or contributors to the pathogenesis of CeD.

Further exploring the interaction between gut microbiota and CeD, González-García et al. (2023) demonstrated bidirectional causality, particularly in individuals carrying the high-risk HLA-DQ2 haplotype. Their findings emphasize the role of specific bacterial families like Ruminococcaceae and Lachnospiraceae, which may both contribute to and result from CeD. Notably, increases in Ruminococcaceae UCG010 and Lachnospiraceae UCG008 significantly raised the odds of CeD, while other bacteria like Anaerotruncus and Tyzzerella3 showed decreased abundance in CeD cases.

Li et al. (2023) provided a comprehensive assessment of how gut microbiota and various metabolites are associated with CeD. Their results identified protective and risk-enhancing roles for different bacterial genera and metabolites. For instance, increased levels of Bifidobacterium and Bifidobacteriales were associated with higher CeD risk (OR = 1.447 and 1.483, respectively), whereas Lentisphaerae, Coprobacter, and Subdoligranulum demonstrated protective effects. Additionally, metabolites like 1-oleoylglycerophosphoethanolamine and 1-palmitoylglycerophosphoethanolamine were linked to increased CeD risk, whereas 10-undecenoate and tyrosine showed protective effects.

Xu et al. (2022) also found a significant association between the abundance of the Bifidobacterium genus and an increased risk of CeD (OR = 1.401, 95% CI = 1.139– 1.722, P_FDR = 2.03 × 10⁻³). Their findings suggest that while Bifidobacterium might typically be considered beneficial, its higher abundance could be detrimental in the context of CeD.

### Neoplastic disease

Four studies evaluated the genetic causal association between CeD and different kinds of neoplastic diseases (**Figures 3,4**).

#### Colorectal and Gastric Cancer

In studies examining gastrointestinal cancers, both Chen et al. (2024) and Wei et al. (2023) found no significant causal relationships between CeD and colorectal or gastric cancer, respectively. Chen et al. reported an odds ratio (OR) of 1.016 (95% CI: 0.983–1.051, P = 0.343) for colorectal cancer, while Wei et al. observed an OR of 0.9509 (95% CI: 0.8416–1.0744, P = 0.419) for gastric cancer.

#### Liver Cancer

Similarly, Yin et al. (2023) explored the potential link between CeD and liver cancer using data from the FinnGen project. Their findings indicated no significant association (OR = 0.91, 95% CI: 0.74–1.11), with consistent results across various Mendelian Randomization (MR) methods and no evidence of heterogeneity or horizontal pleiotropy.

#### Mature T and NK Cell Lymphomas

In contrast to the findings on gastrointestinal and liver cancers, Masot et al. (2023) presented evidence that CeD might significantly increase the risk of mature T and NK cell lymphomas (IVW OR = 1.72, 95% CI: 1.18–2.53, P = 0.00532). This study is particularly notable as it suggests a specific immune-mediated pathway involving T cell activation, highlighted by genetic loci such as TAGAP, which could be implicated in the malignization process of these lymphocyte types.

### Other health comorbidities

The rest of the included studies (n =15), focused on a wide range of health comorbidities, from psychiatric disorders to cardiovascular diseases (**Figures 3,4**). The relationship between celiac disease (CeD) and infectious diseases, particularly during the COVID-19 pandemic, has garnered considerable attention. Zou et al. (2024) demonstrated that genetic predispositions to critical COVID-19 are causally linked to decreased Victivallaceae abundance, subsequently increasing CeD risk (OR = 1.115, 95% CI: 1.007–1.234, P = 0.035). This finding suggests that severe COVID-19 outcomes might elevate CeD risk through alterations in gut microbiota.

Conversely, Li et al. (2022) suggested a protective effect of CeD’s genetic profile against both the susceptibility and severity of COVID-19 (OR = 0.963, 95% CI: 0.937–0.989), indicating potential benefits of CeD genetics against the virus.

Expanding the investigation, Yuan et al. (2024) conducted a phenome-wide study revealing that CeD is associated with increased risks of diseases such as Type 1 diabetes (OR = 1.09, P < 0.01) and systemic lupus erythematosus (OR = 1.10, P < 0.01), along with protective effects against prostate diseases and heightened risks for non-Hodgkin’s lymphoma (OR = 1.03, 95% CI: 1.01-1.05) and osteoporosis (OR = 1.01, 95% CI: 1.00-1.02).

Further research into CeD’s impact on liver and cardiovascular health was explored by Xu et al. (2023) and Huang et al. (2022), respectively. Xu et al. posited CeD as a protective factor against non-alcoholic fatty liver disease (NAFLD) (OR = 0.973, P = 0.026). Meanwhile, Huang et al. found no significant causal relationships between CeD and major cardiovascular outcomes.

Additional studies have examined environmental influences on CeD. Wen et al. (2024) reported that lower exposure to air pollutants like NOX and PM2.5 may decrease the risk of CeD (OR: 0.14, CL: 0.04 - 0.43, OR: 0.17, CL: 0.05 - 0.55). Zhang et al. (2024) addressed the novel link between CeD and facial aging, finding that genetic markers related to CeD slightly increase the risk of facial aging (OR = 1.002, 95% CI: 1.001–1.004, P = 0.009), a relationship that persisted even after adjusting for lifestyle factors.

The studies by Maihofer et al. (2024) and Ruan et al. (2023) explored the intersection between mental health disorders and CeD. Maihofer et al. focused on the effects of genetically predicted PTSD on CeD. Their analysis revealed a significant inverse association, suggesting that individuals with genetic markers for PTSD may have a lower risk of developing CeD (OR = 0.87, 95% CI: 0.78, 0.97). This study implies that PTSD might exert a protective effect against the development of autoimmune diseases like CeD. Conversely, Ruan et al. examined the relationship between genetic liability to depression and a range of gastrointestinal diseases, including CeD. Their findings indicated no significant association between the genetic predisposition to depression and the risk of CeD (OR = 0.64, 95% CI: [0.36, 1.15], p = 0.138).

Research into less common comorbidities, such as Zhou et al. (2024)’s study on frozen shoulder, revealed non-significant associations (OR = 1.02, P = 0.01). Additionally, no significant associations were found between CeD and Amyotrophic Lateral Sclerosis (ALS), senile cataract (**Figure 5**).

**Figure 5:**
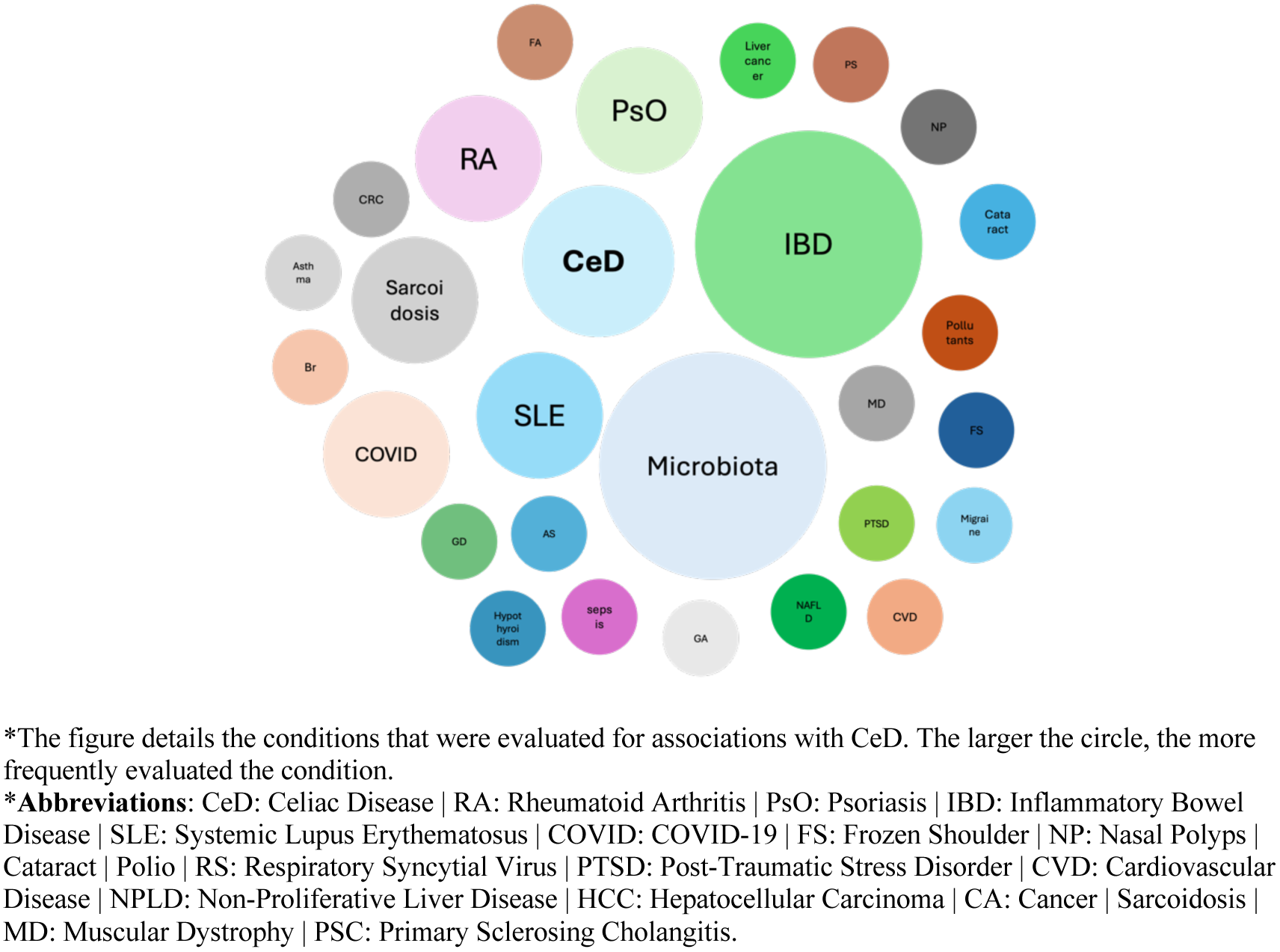
The evaluated associations with their corresponding frequency.

**Figure 5** showcases the frequencies of each studied association.

## Discussion

Our systematic review encompassed 35 studies, predominantly employing robust methodologies. Most of the studies assessed the strength and independence of the instrumental variables from potential confounders and checked and reported horizontal pleiotropy while utilizing sensitivity analyses. The results highlight multiple associations, particularly with autoimmune diseases such as IBD, Type 1 diabetes (T1D), RA, and SLE, alluding to a shared genetic predisposition that may accelerate disease pathogenesis in susceptible individuals. Conversely, the findings indicate protective genetic correlations between CeD and conditions like bronchiectasis and certain metabolic disorders, suggesting a complex interplay of genetic factors. Interestingly, most studies did not find significant associations with various cancers, although one study did identify a significant increase in risk for Mature T/NK cell lymphomas.

When evaluating the robustness of the implied associations from our review, the findings present a nuanced picture. Although many studies reported significant associations, the ORs were frequently close to 1, raising questions about their clinical relevance and the strength of these associations (44). For instance, considering the Region Of Practical Equivalence (ROPE) for binary outcomes—which many included studies address-a suitable ROPE might be considered between 0.83 and 1.17 (45,46). Within this context, several studies reporting significant findings would not be deemed robust. Notably, the associations with psoriasis (OR: 1.12, 95% CI: 1.06– 1.18) and asthma (OR: 1.045, 95% CI: 1.024–1.067) in Li et al.’s study fall into this category. Additionally, the association with facial aging reported by Zhang et al. (OR: 1.002, 95% CI: 1.001–1.004) also sits close to 1, further questioning its impact.

Moreover, several associations reported small ORs or fell well within the ROPE, such as the effects of critical COVID-19 on CeD, CeD’s protective effect against NAFLD, and associations involving various diseases and comorbidities-including sarcoidosis-reported by Yuan et al. Similarly, the protective association of CeD with NAFLD, increased risk of migraine, and the inverse relationship with PTSD show ORs close to 1, highlighting a trend of minimal effect sizes. Interestingly, Sun et al. found a higher risk of sarcoidosis associated with CeD (OR: 1.22), contrasting with Zhou et al.’s lower OR, possibly due to different genetic datasets used, illustrating how data sources can influence outcomes.

These discrepancies across the multitude of associations are well recognized in the Mendelian randomization literature and reflect the inherent variability in such studies (47,48). Nonetheless, the studies consistently distinguished between significant and non-significant results, contributing to a growing understanding of CeD’s complex genetic interactions.

The most consistent and well-studied areas in our review are the association with IBDs and the interactions between CeD and gut microbiota. Regarding IBD, evidence supports a robust, bidirectional relationship with CeD, particularly with Crohn’s disease (CD), as highlighted by studies from Gu et al., Shi et al., and Zhou et al. Specifically, these studies suggest a strong genetic predisposition for CD increases the risk of CeD. These findings align with those from a nationwide register-based cohort study by Marild et al (49). Interestingly, in Marild’s study, the hazard ratio (HR) for CeD in patients with IBD was 5.49 (95% CI 4.90-6.16), with the highest risk estimates observed in ulcerative colitis (HR = 6.99; 95% CI 6.07-8.05), and a lower HR for Crohn’s disease at 3.31 (95% CI 2.69-4.06) (49). Additionally, a systematic review and meta-analysis of 65 case-control studies confirmed a similar association between CeD and IBD (50). However, these results call for a more detailed analysis to clarify the specific connections between CeD and each IBD subtype in future studies (50).

The interaction between CeD and gut microbiota reveals a multifaceted dynamic. Xu et al. and Li et al. report that higher levels of the Bifidobacterium genus and Bifidobacteriales correspond with an elevated risk of CeD, with odds ratios of 1.401 and 1.483, respectively. Conversely, protective effects are observed with the phylum Lentisphaerae, and the genera Coprobacter and Subdoligranulum, suggesting a protective role against CeD.

The complexity extends further as González-García et al. found bidirectional causality between CeD and gut microbiota in individuals with a high-risk HLA-DQ2 haplotype. This study notes significant contributions from Ruminococcaceae and Lachnospiraceae families, indicating potential causal roles for increases in Veillonellaceae and decreases in Pasteurellaceae in relation to CeD development. Additionally, the role of metabolites is crucial, with compounds like 1-oleoylglycerophosphoethanolamine linked to increased CeD risk, whereas 10-undecenoate and tyrosine appear to offer protective effects. Notably, Xu et al.’s reverse analysis found no causal effect of CeD on the abundance of the Bifidobacterium genus, suggesting that while CeD may affect microbiota composition, the influence does not extend universally across all genera. These findings correspond with the current literature and other observational studies (51,52). Nonethelss, they underscore the need for more continued research to explore the intricate connections between CeD, microbiota, and metabolites, aiming to uncover underlying mechanisms and potential therapeutic targets.

Our review synthesized the evidence from a 35 MR studies, and followed a rigorous criteria for evaluating the risk of bias in each included study, following the methodology of a large published systematic review (14). However, this review is not without limitations. The primary constraint is the variability in the quality and size of the cohorts used across the included studies, which may affect the consistency of the findings (48). Moreover, while the review included diverse studies, the majority are from populations of European descent, potentially limiting the applicability of the findings to other ethnic groups. Lastly, the inherent limitations of Mendelian randomization, such as pleiotropy and population stratification, could also influence the results, although efforts were made to mitigate these through careful evaluation and reporting methods.

In conclusion, our study presents a broad spectrum of evidence regarding the causal associations of various diseases with CeD. Key findings include the consistent bi-directional risk increase of IBD in CeD patients and the intricate yet stable associations with changes in gut microbiota. Furthermore, many published results and conclusions about associations are based on weak effect measures, calling into question the significance of these "significant associations." Consequently, further detailed research is necessary to elucidate these associations, especially concerning critical comorbidities such as lymphomas and the genetic predisposition associated with psychiatric morbidity.

## Data Availability

All data produced in the present study are available upon reasonable request to the authors

## Acknowledgment

None

## Contributorship

Mahmud Omar (MO1) - Main contributor, screening, data extraction, writing, validation, and all the steps. Reem Agbareia (RA) – Writing draft, validation, and editing. Saleh Nassar (SN) - Data extraction, validation, and editing. Mohammad Omar (MO2) – screening validation, data extraction and visualization. Mohammad E. Naffaa (MN) - Data validation and editing. Adi Lahat (AL) - Validation and editing. Kassem Sharif (KS) – Screening, data extraction, Validation and oversight.

## Specific Booleans used to screen each data base

### PubMed

("Mendelian Randomization" OR "Mendelian Randomisation" OR "genetic instrumental variables") AND ("Celiac Disease" OR "Coeliac Disease" OR "gluten-sensitive enteropathy")

### OVID (MEDLINE)

("Mendelian Randomization" OR "Mendelian Randomisation" OR "genetic instrumental variables") AND ("Celiac Disease" OR "Coeliac Disease" OR "gluten-sensitive enteropathy")

### Scopus

( TITLE-ABS-KEY ( "Mendelian Randomization" OR "Mendelian Randomisation" OR "genetic instrumental variables" ) AND TITLE-ABS-KEY ( "Celiac Disease" OR "Coeliac Disease" OR "gluten-sensitive enteropathy" ) ) AND ( LIMIT-TO ( DOCTYPE , "ar" ) ) AND ( LIMIT-TO ( EXACTKEYWORD , "Mendelian Randomization Analysis" ) )

### Web of science

(TS=("Mendelian Randomization" OR "Mendelian Randomisation" OR "genetic instrumental variables") AND TS=("Celiac Disease" OR "Coeliac Disease" OR "gluten-sensitive enteropathy"))

### Embase

(’mendelian randomization’/exp OR ’mendelian randomization’ OR ’mendelian randomisation’/exp OR ’mendelian randomisation’ OR ’genetic instrumental variables’) AND (’celiac disease’/exp OR ’celiac disease’ OR ’coeliac disease’/exp OR ’coeliac disease’ OR ’gluten-sensitive enteropathy’/exp OR ’gluten-sensitive enteropathy’) AND [embase]/lim NOT ([embase]/lim AND [medline]/lim) AND ’article’/it

### Cochrane Library

("Mendelian Randomization" OR "Mendelian Randomisation" OR "genetic instrumental variables") AND ("Celiac Disease" OR "Coeliac Disease" OR "gluten-sensitive enteropathy")

