## Supplementary material for "A Systematic Review of Mendelian Randomization Studies on Celiac Disease": Table 1

**Table 1: A summary of the included studies.**

| Author | Year | Exposure Variable | Outcome Variable | Main Results |
|---|---|---|---|---|
| Li et al. | 2023 | CeD (GWAS) | Various autoimmune diseases (GWAS) | Significant ORs: CD (OR: 1.08, 1.04 - 1.11), GD (OR: 1.25, 1.13 - 1.39), PSC (OR: 1.30, 1.23 - 1.39), PsO (OR: 1.12, 1.06 - 1.18), SLE (OR: 1.30, 1.22 - 1.39), T1D (OR: 1.3, 1.23 - 1.38), Asthma (OR: 1.05, 1.02 - 1.07) |
| Zhang et al. | 2024 | CeD (IEU OpenGWAS) | Risk of facial aging (UK Biobank) | OR: 1.002, 1.001 - 1.004 |
| Sun et al. | 2024 | CeD (FinnGen biobank) | Sarcoidosis (FinnGen biobank) | OR: 1.22, 1.15 - 1.28 |
| Zou et al. | 2024 | Critical COVID-19 (COVID-19 Host Genetics Init) | CeD (GWAS) | OR: 1.115, 1.007 - 1.234 |
| Wang et al. | 2024 | 1-oleoylglycerophosphoethanolamine (Metabolomics GWAS server) | CeD (GWAS) | OR: 11.271, 2.053 - 61.882 |
| Su et al. | 2024 | CeD (GWAS) | Bronchiectasis (FinnGen R9 Consortium) | OR: 0.949, 0.902 - 0.999 |
| Zhou et al. | 2024 | CeD (IEU OpenGWAS) | Sarcoidosis (FinnGen consortium, UK Biobank) | OR: OR: 1.13, 1.07 - 1.20 |
| Yuan et al. | 2024 | CeD (GWAS) | Various clinical outcomes (UK Biobank, FinnGen) | Significant ORs: T1D (OR: 1.09, 1.07 - 1.10), GD (OR: 1.06, 1.04 - 1.08), SLE (OR: 1.10, 1.06 - 1.14), Chronic hepatitis (OR: 1.13, 1.08 - 1.18) |
| Chen et al. | 2024 | CeD (GWAS) | Colorectal Cancer (Two GWAS datasets) | OR: 1.016, 0.983 - 1.051 |

| | | | | |
|---|---|---|---|---|
| Li et al. | 2023 | CeD (MRC Integrative Epidemiology Unit, FinnGen) | Sepsis (MRC-IEU, FinnGen) | OR: 1.018, 0.997 - 1.039 |
| Wei et al. | 2023 | CeD (GWAS catalog) | Gastric Cancer (FinnGen database) | OR: 0.9509, 0.8416 - 1.0744 |
| Xu et al. | 2023 | CeD (IEU GWAS) | NAFLD (UK Biobank, Estonian Biobank) | OR: 0.973, 0.949 - 0.997 |
| González-García | 2023 | Gut Microbiota (GWAS of Gut Microbiota by MiBioGen consortium) | CeD (Immunochip CeD study) | Significant ORs: Ruminococcaceae UCG010 (OR: 5.42, exact CI not provided), Lachnospiraceae UCG008 (OR: 4.31, exact CI not provided) |
| Yin et al. | 2023 | CeD (GWAS) | Liver Cancer (FinnGen) | OR: 0.91, 0.74 - 1.11 |
| Li et al. | 2023 | Gut microbiota, Metabolites (MiBioGen consortium, GWAS data of human metabolome) | CeD (GWAS) | Significant ORs: Bifidobacterium (OR: 1.447, 1.054 - 1.988), Bifidobacteriales (OR: 1.483, 1.053 - 2.088) |
| Gu et al. | 2022 | Inflammatory Bowel Disease (GWAS) | CeD (GWAS) | ORs: IBD to CeD (OR: 1.08, 1.03 - 1.14), CeD to CD (OR: 1.04, 1.00 - 1.07) |
| Shi et al. | 2022 | Inflammatory Bowel Disease (IEU GWAS) | CeD (IEU GWAS) | Significant ORs: IBD to CeD (OR: 1.24, 1.16 - 1.34), CD to CeD (OR: 1.27, 1.19 - 1.35) |
| Tang et al. | 2023 | Multisite Chronic Pain (UK Biobank) | CeD (GWAS) | OR: 0.24, 0.02 - 3.64 |
| Masot et al. | 2023 | CeD (GWAS) | Mature T/NK cell lymphomas (FinnGen) | OR: 1.72, 1.18 - 2.53 |
| Yuan et al. | 2023 | CeD (GWAS) | Inflammatory Bowel Disease (GWAS) | OR for CD: 1.1408, 1.0614 - 1.2261 |
| Li et al. | 2022 | CeD (UK Biobank, GWAS) | COVID-19 severity (COVID-19 Host Genetics Initiative GWAS) | OR: 0.919, 0.844 - 0.999 |

| | | | | |
|---|---|---|---|---|
| Xu et al. | 2022 | Gut Microbiota (GWAS) | CeD (GWAS) | OR: 1.401, 1.139 - 1.722 |
| Welander et al. | 2023 | CeD (GWAS) | Migraine (GWAS) | Significant ORs: MO (OR: 0.95, 0.92 - 0.99), MA (OR: 1.04, 1.00 - 1.08) |
| Alipour et al. | 2022 | CeD (IEU OpenGWAS) | Amyotrophic Lateral Sclerosis (MinE GWAS) | OR: 1.016, 0.992 - 1.040 |
| Maihofer et al. | 2024 | Genetically predicted PTSD (PGC-PTSD Freeze 3 GWAS) | CeD (NHGRI-EBI GWAS) | OR: 0.87, 0.78 - 0.97 |
| Ruan et al. | 2023 | Depression (GWAS, UK Biobank, 23andMe, Psychiatric Genomics Consortium) | CeD (UK Biobank, FinnGen) | OR: 0.64, 0.36 - 1.15 |
| Li et al. | 2022 | CeD (GWAS) | Psoriasis (GWAS) | OR: 1.232, 1.061 - 1.432 |
| Huang et al. | 2022 | CeD (GWAS) | Cardiovascular Diseases (IEU GWAS) | Non-significant associations |
| Wang et al. | 2023 | Systemic Lupus Erythematosus (MRC-IEU) | CeD ( MRC-IEU) | OR: 1.24, 1.03 - 1.49 |
| Hua et al. | 2022 | Rheumatoid Arthritis (GWAS) | CeD (GWAS) | OR: 1.46, 1.19 - 1.79 |
| Chen et al. | 2023 | CeD (GWAS) | Nasal Polyps (UK Biobank) | OR: 1.000494, 1.000067 - 1.000922 |
| Yuan et al. | 2024 | CeD (GWASs) | Senile Cataract (FinnGen) | OR: 1.04, 1.01 - 1.08 |
| Zhou et al. | 2024 | Crohn's Disease (GWAS) | CeD (GWAS) | OR: 1.14, 1.03 - 1.25 |
| Wen et al. | 2024 | Air Pollutants (UK Biobank) | CeD ( FinnGen) | ORs: NOX (OR: 0.14, 0.04 - 0.43), PM2.5 (OR: 0.17, 0.05 - 0.55) |
| Zhou et al. | 2024 | CeD (FinnGen) | Frozen Shoulder (UK Biobank) | OR: 1.02, 1.01 - 1.04 |

**Abbreviations**: CeD: Celiac Disease | CD: Crohn's Disease | GD: Graves' Disease | PSC: Primary Sclerosing Cholangitis | PsO: Psoriasis | SLE: Systemic Lupus Erythematosus | T1D: Type 1 Diabetes | GWAS: Genome-Wide Association Studies | IVW: Inverse Variance Weighted | WM: Weighted Median | MO: Migraine without Aura | MA: Migraine with Aura | IBD: Inflammatory Bowel Disease | UC: Ulcerative Colitis | NAFLD: Non-Alcoholic Fatty Liver Disease | MRC-IEU: MRC Integrative Epidemiology Unit | FinnGen: FinnGen Research Project | OR: Odds Ratio | CI: Confidence Interval.
