## Supplement for "A Systematic Review of Mendelian Randomization Studies on Celiac Disease"

**Specific Booleans used to screen each data base:**

**PubMed**

("Mendelian Randomization" OR "Mendelian Randomisation" OR "genetic instrumental variables") AND ("Celiac Disease" OR "Coeliac Disease" OR "gluten-sensitive enteropathy")

**OVID (MEDLINE)**

("Mendelian Randomization" OR "Mendelian Randomisation" OR "genetic instrumental variables") AND ("Celiac Disease" OR "Coeliac Disease" OR "gluten-sensitive enteropathy")

**Scopus**

( TITLE-ABS-KEY ( "Mendelian Randomization" OR "Mendelian Randomisation" OR "genetic instrumental variables" ) AND TITLE-ABS-KEY ( "Celiac Disease" OR "Coeliac Disease" OR "gluten-sensitive enteropathy" ) ) AND ( LIMIT-TO ( DOCTYPE , "ar" ) ) AND ( LIMIT-TO ( EXACTKEYWORD , "Mendelian Randomization Analysis" ) )

**Web of science**

(TS=("Mendelian Randomization" OR "Mendelian Randomisation" OR "genetic instrumental variables") AND TS=("Celiac Disease" OR "Coeliac Disease" OR "gluten-sensitive enteropathy"))

**Embase**

('mendelian randomization'/exp OR 'mendelian randomization' OR 'mendelian randomisation'/exp OR 'mendelian randomisation' OR 'genetic instrumental variables') AND ('celiac disease'/exp OR 'celiac disease' OR 'coeliac disease'/exp

OR 'coeliac disease' OR 'gluten-sensitive enteropathy'/exp OR 'gluten-sensitive enteropathy')

AND

[embase]/lim NOT ([embase]/lim AND [medline]/lim) AND 'article'/it

**Cochrane Library**

("Mendelian Randomization" OR "Mendelian Randomisation" OR "genetic instrumental variables") AND ("Celiac Disease" OR "Coeliac Disease" OR "gluten-sensitive enteropathy")

**Table S1: Risk of bias evaluation of the included studies.**

| Author | IV1 | IV2 | IV3 |
|---|---|---|---|
| **Li et al** | high | moderate | high |
| **Zhang et al** | high | high | high |
| **Sun et al** | high | moderate | high |
| **Zou et al** | high | high | high |
| **Wang et al** | high | high | high |
| **Su et al** | high | high | high |
| **Zhou et al** | high | moderate | high |
| **Yuan et al** | high | high | high |
| **Chen et al** | high | high | moderate |
| **Li et al** | moderate | moderate | poor |
| **Wei et al.** | Moderate | Poor | Poor |
| **Xu et al.** | High | Moderate | High |
| **González-García et al.** | Moderate | High | High |
| **Yin et al.** | High | High | High |
| **Li et al. (3)** | Moderate | High | High |
| **Gu et al.** | High | High | High |
| **Shi et al.** | High | High | High |
| **Tang et al.** | High | Poor | Poor |
| **Masot et al.** | High | High | High |
| **Yuan et al. (2)** | High | High | High |
| **Li et al.** | High | Moderate | High |
| **Xu et al.** | High | High | High |
| **Welander et al.** | High | High | High |
| **Alipour et al.** | High | High | High |

| | | | |
|---|---|---|---|
| **Maihofer et al.** | High | High | High |
| **Ruan et al.** | High | High | High |
| **Li et al. (5)** | High | High | High |
| **Huang et al.** | High | High | High |
| **Wang et al.** | High | High | Moderate |
| **Hua et al** | High | High | High |
| **Chen et al.** | High | High | High |
| **Yuan et al.** | High | High | High |
| **Zhou et al. (2)** | High | High | High |
| **Wen et al.** | High | High | High |
| **Zhou et al. (3)** | High | High | High |

**Abbreviations**: IV: Instrumental variable | IV1: Relevance | IV2: Independence | IV3: Exclusion-restriction.
